# Synthetic Data Generation in Motion Analysis: A Generative Deep Learning Framework

**DOI:** 10.1101/2024.09.27.24314497

**Authors:** Mattia Perrone, Steven Mell, John Martin, Shane J. Nho, Scott Simmons, Philip Malloy

**Affiliations:** Section of Young Adult Hip Surgery, Division of Sports Medicine, Department of Orthopedic Surgery, Rush Medical College of Rush University, Rush University MedicalCenter, Chicago, Illinois, USA; Department of Mathematics and Computer Science, Drury University, Springfield, Missouri, USA; Department of Physical Therapy, Arcadia University, Glenside, Pennsylvania, USA

## Abstract

Generative deep learning has emerged as a promising data augmentation technique in recent years. This approach becomes particularly valuable in areas such as motion analysis, where it is challenging to collect substantial amounts of data. The current study introduces a data augmentation strategy that relies on a variational autoencoder to generate synthetic data of kinetic and kinematic variables. The kinematic and kinetic variables consist of hip and knee joint angles and moments, respectively, in both sagittal and frontal plane, and ground reaction forces. Statistical parametric mapping (SPM) did not detect significant differences between real and synthetic data for each of the biomechanical variables considered. To further evaluate the effectiveness of this approach, a long-short term model (LSTM) was trained both only on real data (R) and on the combination of real and synthetic data (R&S); the performance of each of these two trained models was then assessed on real test data unseen during training. The predictive model achieved comparable results in terms of nRMSE when predicting knee joint moments in the frontal (R&S: 9.86% vs R:10.72%) and sagittal plane (R&S: 9.21% vs R: 9.75%), and hip joint moments in the frontal (R&S: 16.93% vs R:16.79%) and sagittal plane (R&S: 13.29% vs R:14.60%). These findings suggest that the proposed methodology is an effective data augmentation approach in motion analysis settings.

## 1. Introduction

The applications of motion analysis have expanded over the years (1), being employed in a wide range of fields from sports science (2) to robotics (3) and computer animation (4). Biomechanics continues to rank among the most prominent areas for applications of human motion analysis, where this technology is used to measure kinetic and kinematic variables of participants performing various functional activities (5,6, 7,8). Such motion analysis is often employed to gauge the progression of disease or to understand healthy movement in the context of optimal performance (9, 10,11,12,13,14). A primary challenge in human motion capture research stems from the significant time commitment inherent in participating to a study and the resulting difficulty in recruiting participants. Additionally, the financial resources required to perform a large scale motion capture study are considerable, leading to small samples sizes in most motion capture investigations.

The popularity of machine learning has grown steadily in the field of motion capture biomechanics (15,16,17 , 18 , 19 ). Most of these investigations focus on the prediction of biomechanical variables during specific motion tasks (20,21,22,23,24). A major constraint when deploying machine learning techniques in motion capture biomechanics is the limited amount of data that can be collected to fit a model (20). Various strategies have been employed to augment motion capture datasets (25,26), but without fully resolving the issue of generating synthetic data that faithfully resemble real data.

Generative artificial intelligence (AI) has gained wide application in the past few years, particularly in the realm of image (27) and language (28) generation. In the field of motion capture biomechanics, generative AI can be employed to augment datasets by generating synthetic biomechanical variables such as joint angles and joint moments. This technique entails creating new data using a generative AI model, thus increasing the size of the dataset without collecting new data from the real world. However, the number of studies in motion capture biomechanics involving generative AI is still small, likely due to the limited availability of generative model architectures that can effectively handle time series data as input. A few studies employed a generative adversarial network (GAN) to augment motion capture data previously collected in laboratory settings (29,30) Although these investigations included only a small number of subjects, they showed that generative deep learning can be successfully employed for data augmentation purposes in motion analysis. The aim of the current paper is to introduce both a deep learning (DL) approach for data augmentation and a DL based application to evaluate its effectiveness. Synthetic trials were generated from real trial data using a variational autoencoder (VAE), a DL architecture that is easier and faster to train with respect to other generative DL models, making it more suitable when the training set includes a limited number of observations (31). Statistical parametric mapping (SPM) between real and synthetic data was performed to assess the quality of synthetic data. To further gauge the quality and utility of synthetic data, the performance of a long short-term memory (LSTM) model (32) trained on the original dataset has been compared to the performance of the same predictive model trained on both real and synthetic data. To assess the predictive LSTM model performance, nRMSE for hip and knee moments was calculated in both scenarios (real data and real combined with synthetic data) using the predictions from musculoskeletal (MSK) models as gold standard. nRMSE was defined as the square root of the average of the squared differences between the predicted and observed values normalized by the range of observed values.

## 2. Methods

### 2.1. Study Design and Data Acquisition

The data included in this study were selected from a larger longitudinal motion capture study that investigated participants with hip conditions and healthy controls (11,33). A total of 53 subjects, comprising 24 healthy controls and 29 patients affected by femoroacetabular impingement syndrome, provided written informed consent based on local institutional review board standard and participated in the study. Each participant performed 6 single leg squat trials (3 for each leg), for a total of 318 observations. A total of 115 observations were selected out of the 144 for healthy controls; the remaining 29 observations were discarded because of dropout markers and/or mismatch between biomechanical variables. To avoid bias when training the model, a total of 115 observations were selected as well among the 174 available for patients. Therefore, the final dataset includes 230 samples. Single leg squat was chosen as motion task since individuals with femoroacetabular impingement syndrome exhibit distinct patterns in biomechanical variables compared to healthy subjects during this task, such as lower hip flexion angles and lower hip flexion moments (14,32,34). Moreover, single leg squat has been recommended as valid and reliable task for patients with non-arthritic hip pain and is commonly used in functional clinical assessments for these patients (35,36,37).

### 2.2. Data Processing with Visual3D

All the motion capture data collected were exported from the data acquisition software (Motive, Natural Point Inc.) to a commercially available biomechanics analysis software (Visual3D, C-Motion), as described elsewhere (32). The biomechanical variables processed and exported from Visual3D were hip joint angles (HJA), hip joint moments (HJM), knee joint angles (KJA), knee joint moments (KJM) in the sagittal and frontal plane, and vertical ground reaction forces (GRF). These values represent the gold standard in the current study.

### 2.3. Data Augmentation using Variational Autoencoder

Trials were cropped to single leg squat cycles and time-normalized to 100 frames each. Each observation in the dataset consisted of 9 time series, corresponding to the 9 biomechanical variables processed in Visual3D: HJA, HJM, KJA, KJM in both the sagittal and frontal planes, as well as GRF. The shape of the dataset is therefore 230×100×9. Each input feature was separately normalized to fall within the range of 0 to 1 to help model predictions, as standard practice (29). Data augmentation has been performed using a variational autoencoder (figure 1). At first, we attempted to use generative adversarial networks (GAN) for data augmentation (38); however, due to our limited dataset, we were unable to achieve convergence and therefore opted to use a VAE. The VAE architecture has first been proposed by Kingma et al. (31) to generate realistic samples starting from multivariate time series (39,40). Autoencoders are a type of neural network that learns to encode and decode data, reconstructing the input data by learning a compressed representation of it (41). It comprises two primary components: an encoder network, responsible for mapping input data into a lower-dimensional latent space, and a decoder network, which reconstructs the original data space from the latent representation. The encoder and decoder networks are trained simultaneously using a loss function that encourages the encoder to produce latent variables capturing the underlying structure of the data, and the decoder to generate data that are similar to the original input. The key innovation of variational autoencoders (VAE) with respect to autoencoders is the use of probabilistic inference in the latent space, which allows the model to generate new data starting from the parameters (mean and standard deviation) of the latent distribution (31). Since the input data consist of multivariate time series, long term-short memory (LSTM) layers were included in both the encoder and the decoder architecture to capture temporal dependencies within the data more effectively. LSTMs integrate current data with both short-term and long-term ‘memory’ to enable the learning of time-global patterns during training. In the context of our VAE architecture, this global information is both encoded and decoded so that synthetic trials may represent the time development of real trials. Finally, grid search hyperparameter tuning was performed, considering as hyperparameters the first and last hidden layer size, respectively for the encoder and the decoder (128, 256 and 512), the latent vector length (32, 64 and 128), and the learning rate (0.01 and 0.001). The best hyperparameter combination uses 512 hidden units with a latent vector length of 128 and a learning rate of 0.001. The results of hyperparameter tuning were assessed using 10- fold cross-validation, with mean squared error (MSE) as loss function. The model with the best hyperparameters was trained for 1000 epochs on the training set, which consists of a randomly selected 75% of the total number of samples (n = 172). The testing dataset was isolated during the training of the generative model to avoid any data leakage. Initially, we split the 230 samples into 172 for training (86 controls and 86 patients) and 58 for testing (29 controls and 29 patients). The 172 training samples were then augmented separately by cohort to a total of 430 samples. This means the autoencoder was trained from scratch twice: once using 86 controls (generating 129 synthetic controls) and once using 86 patients (generating 129 synthetic patients). In both scenarios, the number of samples generated is 1.5 times the real data, meaning that the final dataset underwent a 2.5- fold augmentation compared to the initial one. This parameter was determined by referencing another study focused on generative AI in motion analysis settings (29). Finally, the predictive model was trained on these 430 observations and tested on the 58 samples isolated at the beginning.

**Figure 1.**
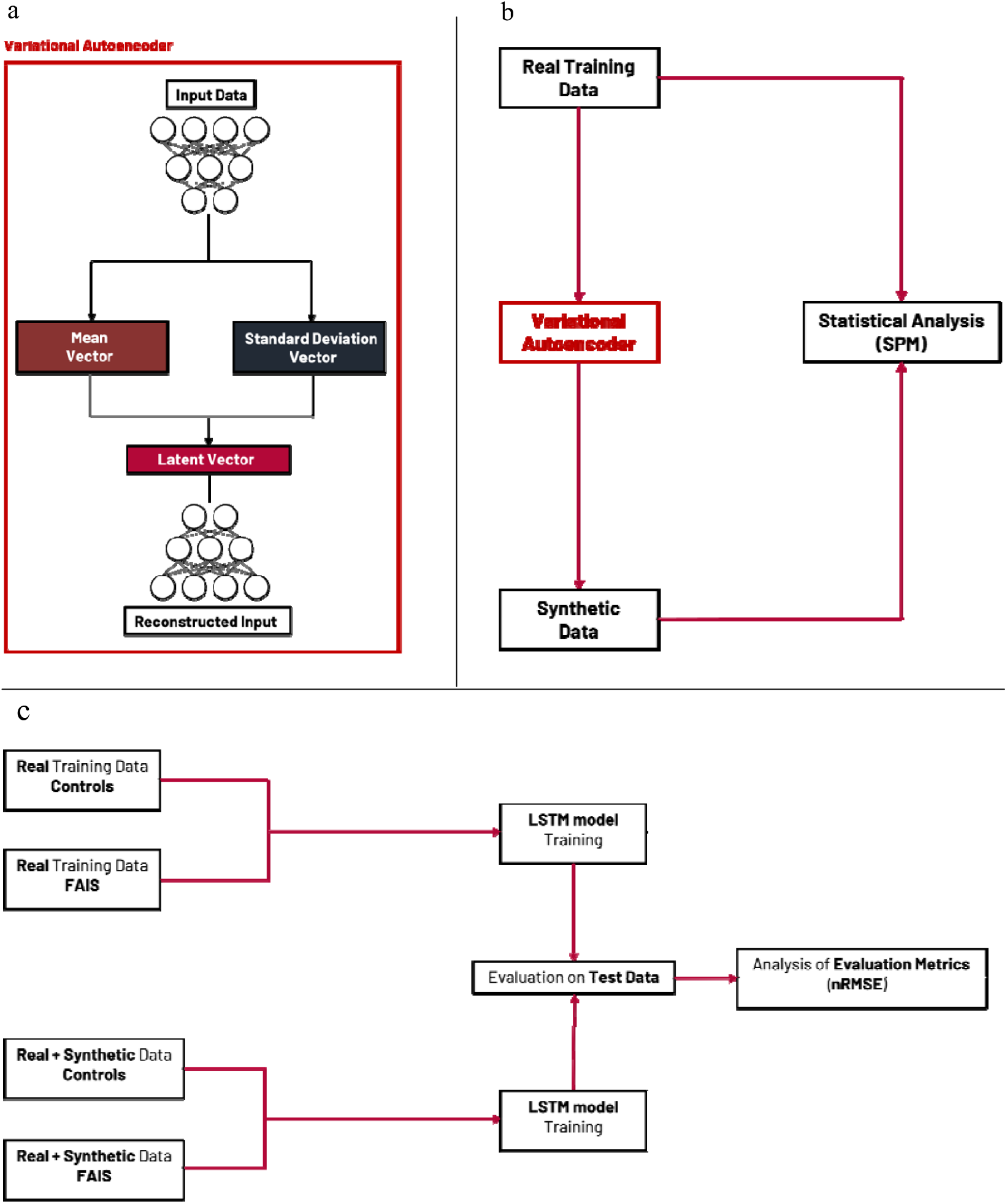
(a) Schematic of the variational autoencoder architecture. (b) Pipeline followed to generate synthetic data and assess their quality. This was done separately for patients and controls. (c) Pipeline followed to compare performances of the predictive LSTM model trained only on real data and on real and synthetic data.

### 2.4. LSTM model training

To assess the effectiveness of this data generation process, a further analysis has been conducted. This consists in training an LSTM predictive model, already introduced elsewhere (32), both on the real and on the combination of real and synthetic data, comparing performance in these two scenarios. The dataset used to train the predictive model comprises both controls and patients. The input data were GRF, HJA and KJA in sagittal and frontal plane. The model was trained 4 times, considering as output variables sagittal plane HJM, frontal plane HJM, sagittal plane KJM and frontal plane KJM. Root mean square error normalized to the range of the data (nRMSE) was calculated between the gold standard, which are the kinetic variables calculated through Visual3D, and the predictions from both real data and the combination of real and synthetic data, to evaluate model performances. Data processing and implementation of both DL models (VAE and LSTM) were carried out in Python 3 utilizing the Pandas, Numpy, Scipy, Scikit learn, Keras and Tensorflow frameworks. Both VAE and LSTM training were carried out on a NVIDIA T4 GPU.

### 2.5. Statistical Analysis

The synthetic data generated by the VAE were compared with the gold standard using statistical parametric mapping (SPM) in Python (42). SPM was performed between real and VAE-generated data separately for controls and patients. An unpaired samples t-test was performed and an alpha level of 0.05 was used to define statistical significance.

## 3. Results

### 3.1. Variational Autoencoder

The best hyperparameter configuration of the VAE includes 512 hidden units, a latent vector length of 128 and a learning rate of 0.001. Using these parameters, the VAE generated kinetic and kinematic variables for both controls (figure 2) and patients (figure 3).

**Figure 2.**
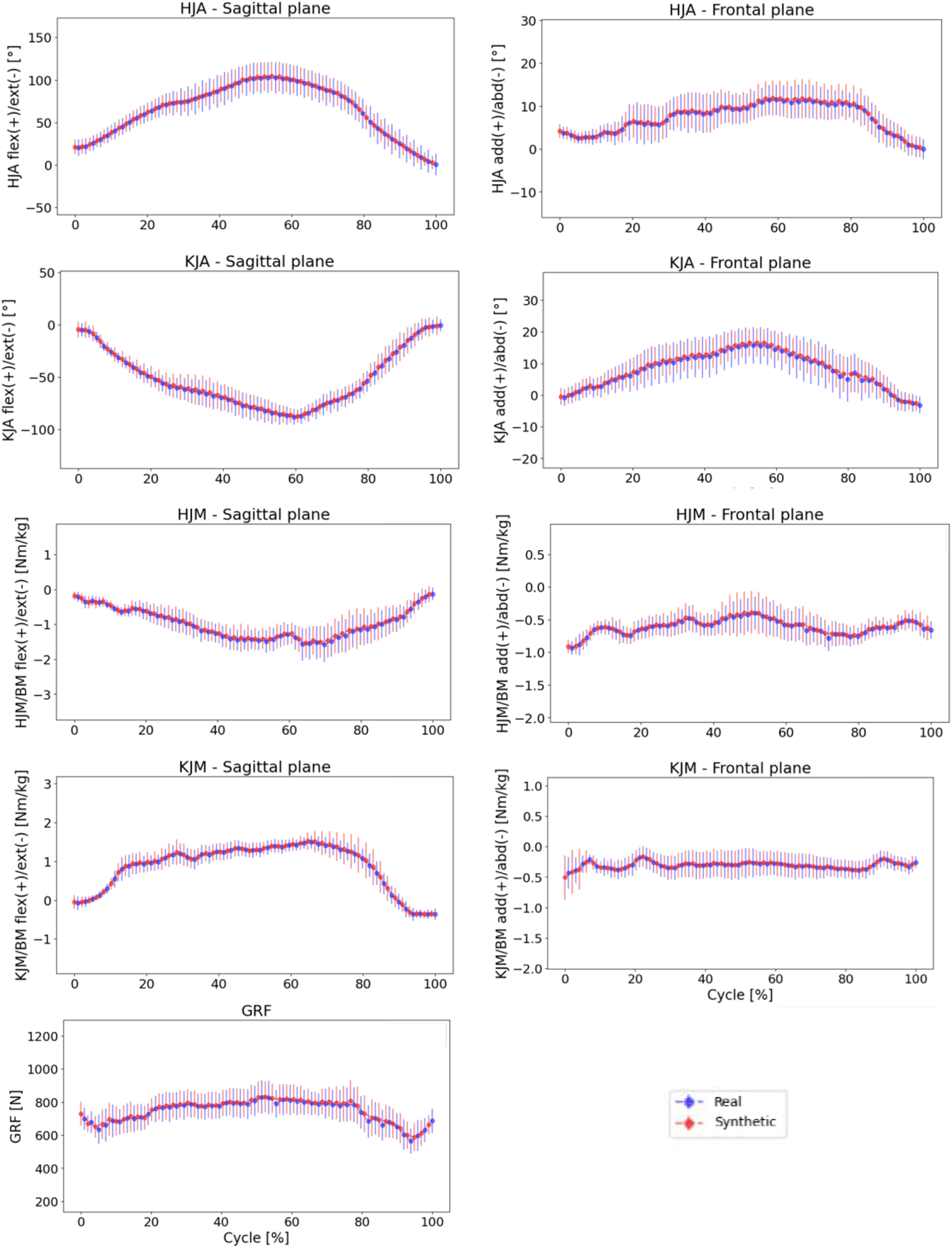
Graphs showing waveforms of all the biomechanical variables analyzed for controls. Dots show the mean of the variables of interest and error bars represent standard error of real (blue) and synthetic (red) data. No statistically significant differences are observed for any variable.

**Figure 3.**
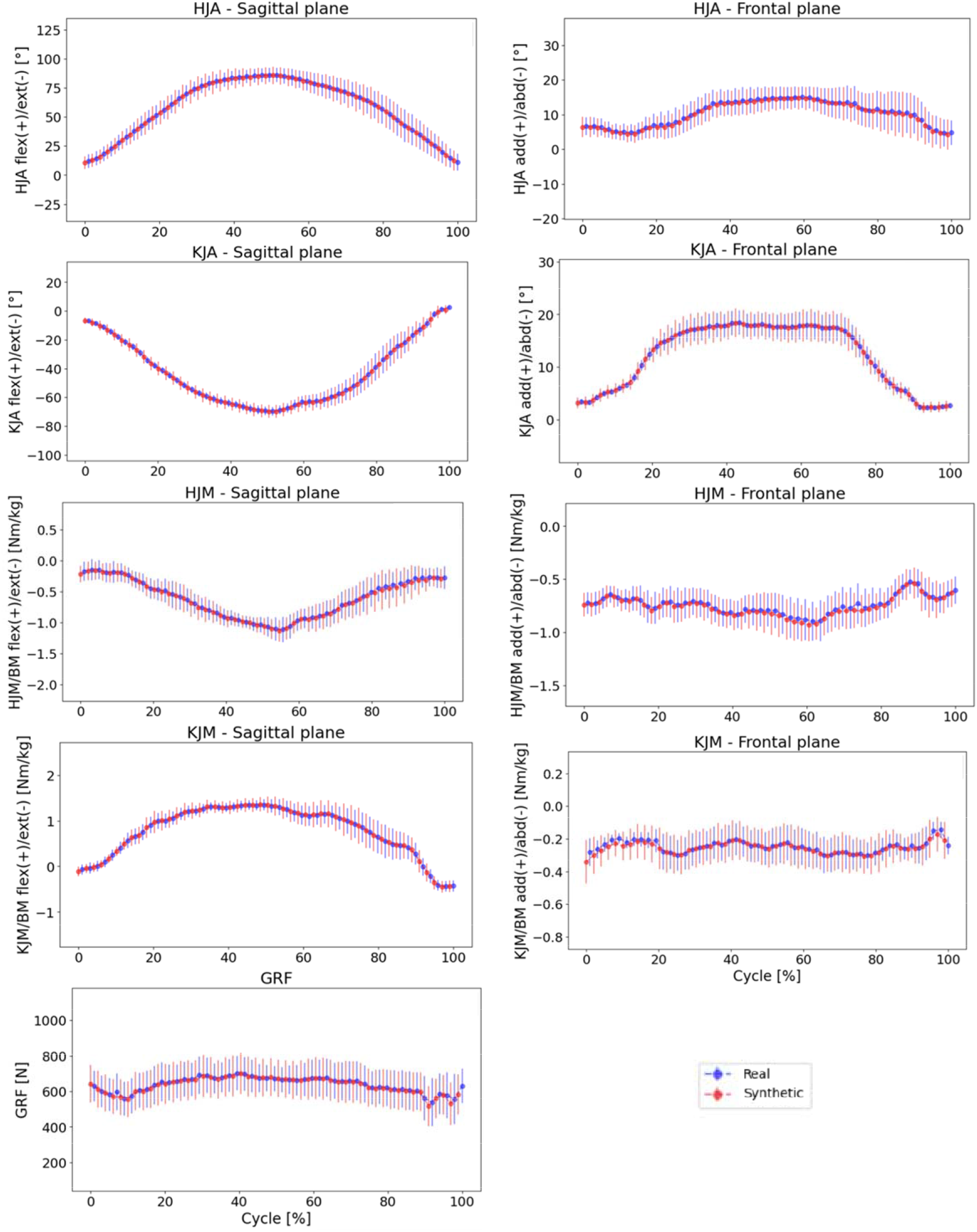
Graphs showing waveforms of all the biomechanical variables analyzed for patients. Dots show the mean of the variables of interest and error bars represent standard error of real (blue) and synthetic (red) data. No statistically significant differences are observed for any variable.

3.2. LSTM model

An LSTM model (32) was used in the current study to make predictions of flexion and abduction moments at the hip and knee. Input data were GRF, flexion and abduction angles at the hip and knee. The model trained on a combination of real and synthetic data demonstrated performance comparable to that of the model trained solely on real data (figure 4 and figure 5), as also confirmed by nRMSE values for both models (Table 1).

**Table 1:**
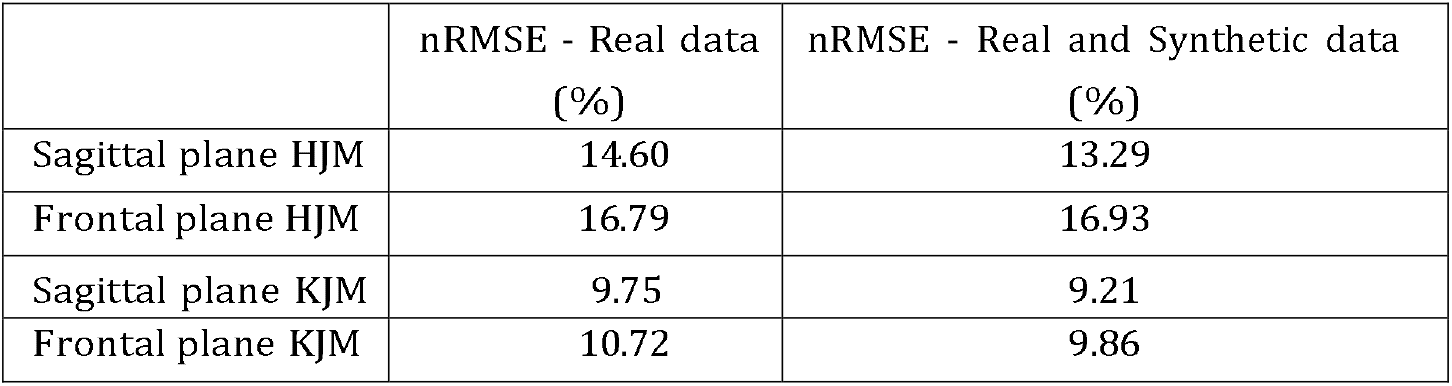
nRMSE between test data and predictions of the LSTM model trained on only real data (first column) and between test data and predictions of the LSTM model trained on both real and synthetic data (second column).

**Figure 4.**
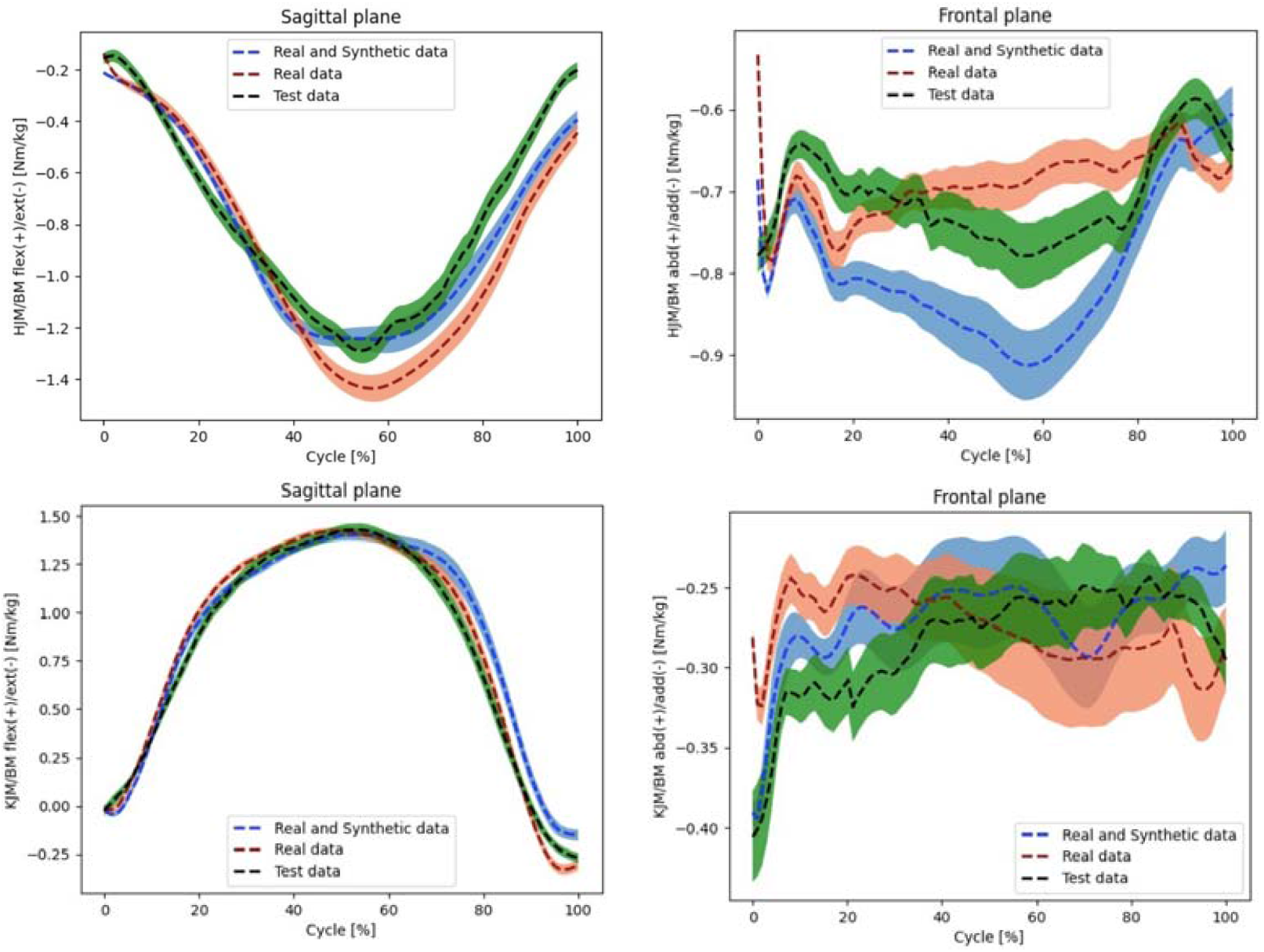
Mean and standard error of the 4 output biomechanical variables predicted by the LSTM model (flexion and adduction HJM and KJM). Green, red and blue curves represent respectively gold standard test data, predictions of the model trained on only real data and predictions of the model trained on both real and synthetic data, with the respective standard errors.

**Figure 5.**
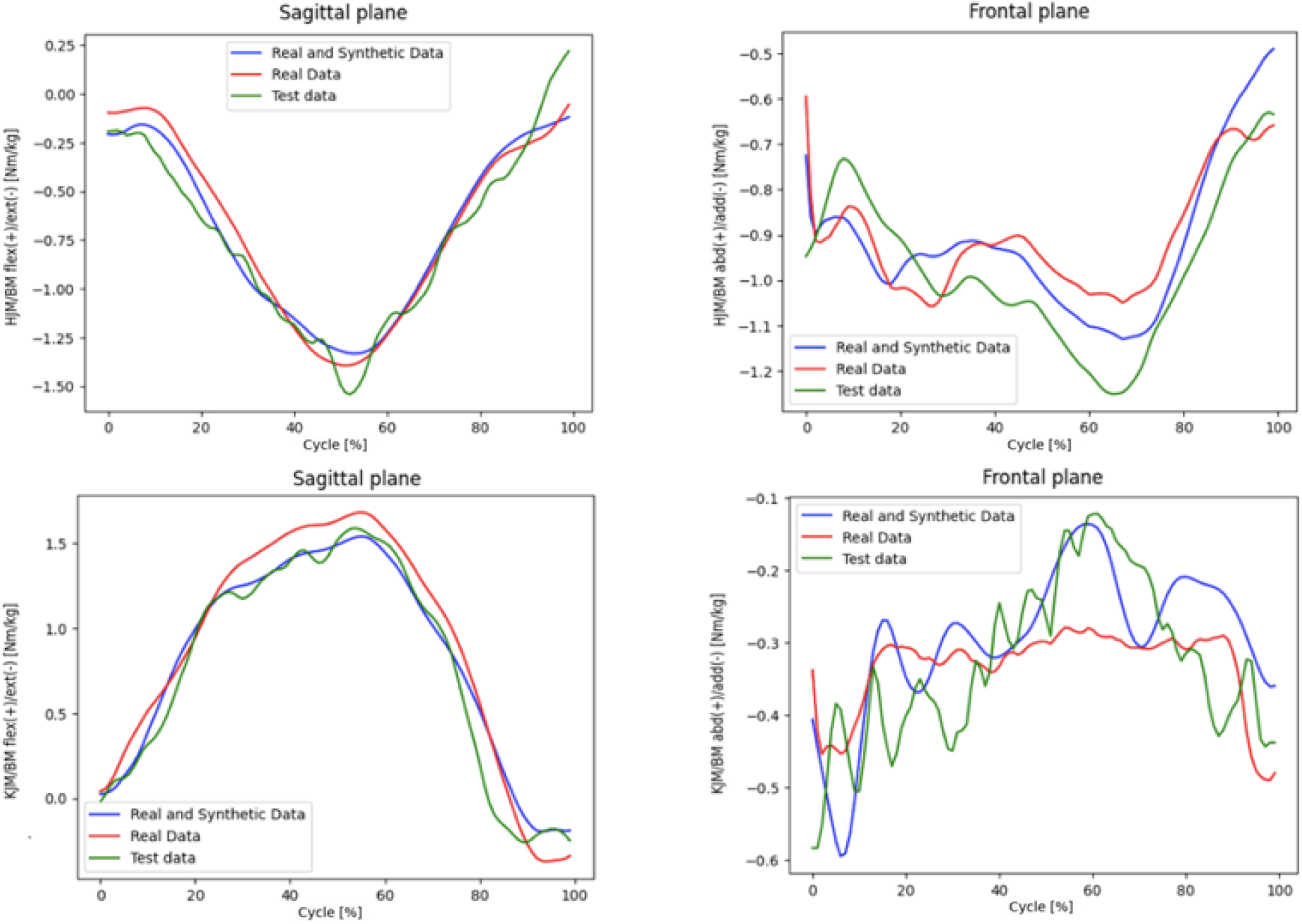
Comparison of a test set sample for the 4 output biomechanical variables predicted by the LSTM model (flexion and adduction HJM and KJM). Green, red and blue curves represent respectively gold standard test data, predictions of the model trained on only real data and predictions of the model trained on both real and synthetic data.

## 4. Discussion

### 4.1. Variational Autoencoder

No differences were found between real and synthetic data for any of the 9 variables from SPM both when considering healthy controls and patients. An aspect to consider when assessing the quality of synthetic data is their standard deviation or standard error. If synthetic data have a lower standard deviation than real data, this means that the generated data are not able to capture the full range of variability of the original real data. On the other hand, excessively high variability in synthetic data could raise concerns about the reliability and consistency of the generated samples. The dataset ’s small size could be a contributing factor of these differences, making it challenging to generate synthetic data that accurately represent the entire original population (figure A.6 and figure A.7).

### 4.2. LSTM model

The model trained on real and synthetic data achieved comparable performance in terms of nRMSE when predicting sagittal plane and frontal plane HJM and KJM (table 1). It is important to highlight that while the incorporation of synthetic data does not worsen the LSTM model performance, only modest improvements are observed (table 1). A possible explanation for this could be attributed to the limited amount of training data available for the variational autoencoder (VAE). Data augmentation, while a powerful tool in machine learning, shows its limitations when the amount of original data is constrained. Studies have shown that the effectiveness of data augmentation strategies based on generative approaches tends to be less significant in scenarios where the dataset size is limited (43). On the other hand, it is worth noting that motion analysis datasets are typically smaller relative to those in other fields of research. With 230 samples, the current study falls in the top 15% for sample size of studies dealing with machine learning in human movement biomechanics as reported by Halilaj et al (20).

### 4.3. Comparison with literature

One of the main challenges faced by researchers when conducting motion analysis studies is overcoming limitations in the amount of data collected in each trial. Data augmentation can be employed to address this issue and generative AI stands out as one of the most effective approaches for dataset expansion (44). Due to the limited number of observations in these kinds of investigations, VAEs offer a suitable modeling approach. In the current study, the performance of an LSTM model trained on both real and synthetic data has been compared to the performance of the same model trained just on real data, to demonstrate the effectiveness of the proposed data augmentation approach. Each sample of the dataset consists of 9 biomechanical time series of subjects performing single leg squats: GRF, hip and knee joint angles and moments in both the sagittal and the frontal plane. This study encompassed both healthy controls and patients with musculoskeletal conditions, to demonstrate the generalizability of the current model across diverse populations.

The current study is one of the first dealing with generative AI in motion analysis. Another study leverages VAE for data augmentation of motion capture datasets, but with an emphasis on enhancing general human motion prediction, rather than focusing on synthetic biomechanical variables generation (45). A few studies employed a generative adversarial network (GAN) to augment motion capture datasets by generating synthetic biomechanical variables (29,30). The current investigation presents some novelties with respect to these studies. First of all, it features a substantially larger number of participants (n=53) with respect to the sizes of the groups in the previous studies (respectively n=8 and n=5). This increased subject count enhances the diversity of the test set, strengthening the overall reliability of the results. Additionally, our study encompasses different cohorts, including both healthy controls and patients diagnosed with a hip condition (femoroacetabular impingement syndrome), demonstrating the robustness of our framework across varied populations. Finally, we applied this framework for the first time to a motion task different than gait, specifically focusing on the single leg squat.

However, it remains challenging to compare the VAE performance with that in the literature, both due to the limited number of similar studies and because the quality of the synthetic data highly depends on the quality of the real data. When comparing real and synthetic data, no statistically significant differences were found by SPM for each of the variables in the dataset. To assess the performance of the prediction model, nRMSE was used as the evaluation metric. The use of nRMSE allows us to compare results of the current study with those in the literature that analyze different motion tasks and/or biomechanical variables. This error metric ranges from 10% to 20% in other investigations dealing with machine learning in biomechanics, (25,46,47) which is consistent with our findings (Table 1).

### 4.4. Limitations

The current study has some limitations. The size of the dataset utilized in this study is relatively small by machine learning standards; it is likely that leveraging larger datasets would lead to improved performance for both the VAE and LSTM models employed in this study. Moreover, the investigation does not address specifically how the amount of synthetic data in the training set affects the LSTM model performance. Finally, the study does not explore how the number of samples used to train the VAE influences the quality of synthetic data and, consequently, the predictive model’s performance.

## 5. Conclusion

The current study proposes a novel approach for data augmentation of motion capture data using generative deep learning. A variational autoencoder has been used to generate synthetic samples. To assess the quality of synthetic data, the performances of an LSTM predictive model trained both only on real data and on the combination between real and synthetic data have been compared. The model achieved comparable nRMSE on the

prediction of all 4 biomechanical outputs when trained on both real and synthetic data. Investigating the impact of synthetic data volume on the predictive model performance, as well as exploring the influence of the generative model training sample size on the quality of synthetic data, could be valuable paths for future research.

## Data Availability

All data produced in the present study are available upon reasonable request to the authors

## Acknowledgments

The authors acknowledge the Michael and Jacqueline Newman orthopaedic research fund for the support of this work.

## Declaration of Conflicting Interests

We disclose the following conflicts of interest: Mattia Perrone (none), Steven P. Mell (none), John Martin (none), Scott Simmons (none), Philip Malloy (none), Shane J. Nho (Allosource: Research support, American Orthopaedic Association: Board or committee member, American Orthopaedic Society for Sports Medicine: Board or committee member, Arthrex, Inc: Research support, Arthroscopy of North America: Board or committee member, Athletico: Research support, DJ Orthopaedics: Research support, Linvatec: Research support, Miomed: Research support, Ossur: IP royalties, Smith Nephew: Research support, Springer: Publishing royalties, financial or material support, Stryker: IP royalties; Paid consultant; Research support).

## Notes

### Funding Statement

No funding

### Author Declarations

The IRB of Rush University gave full ethical approval for this study

