## Supplementary material for "Synthetic Data Generation in Motion Analysis: A Generative Deep Learning Framework"

Appendix A. Supplementary Material

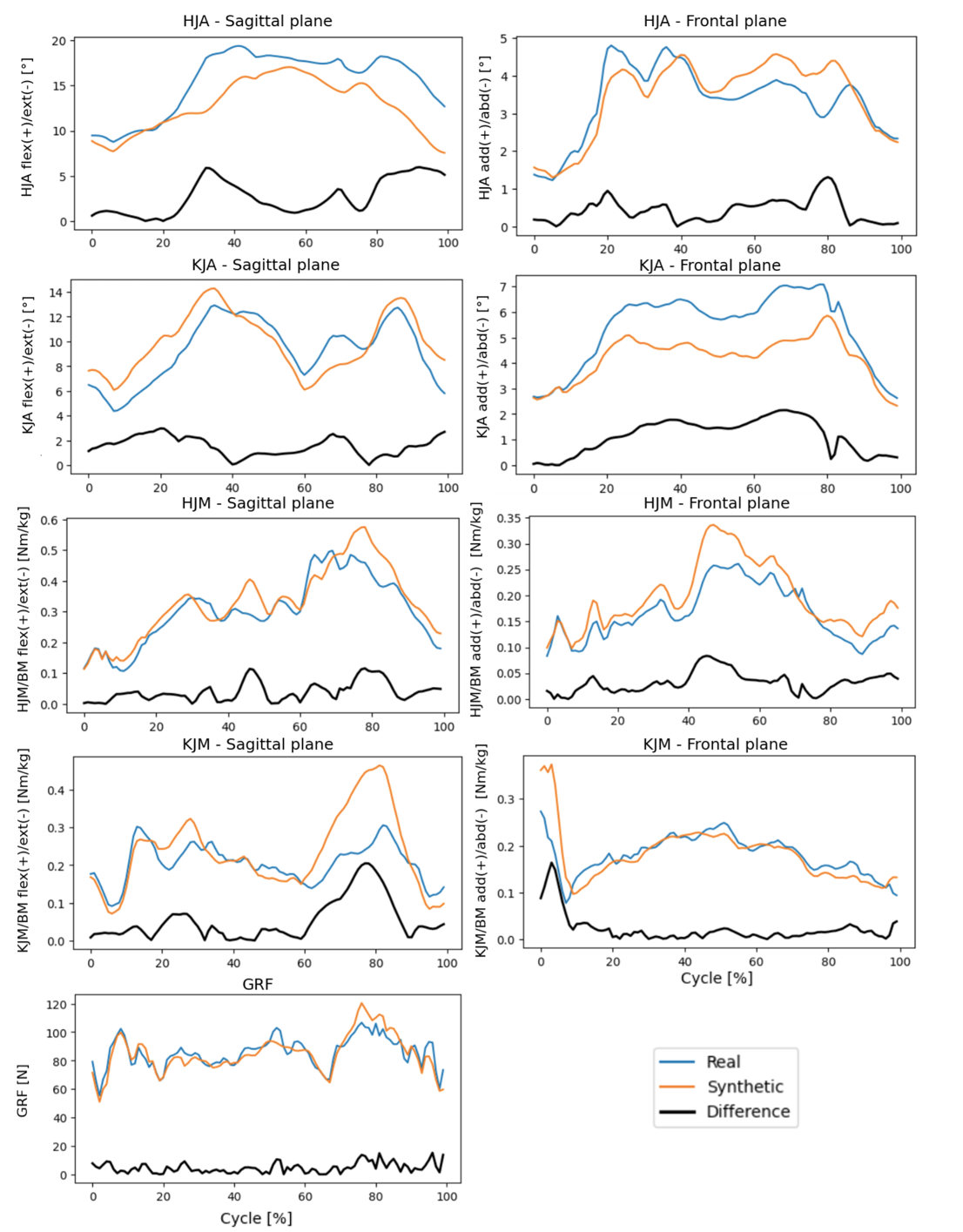

Figure A.6: Graphs showing standard deviation waveforms of all the biomechanical vari- ables analyzed for controls. At each time step, standard deviations were calculated across all corresponding real (blue) and synthetic (orange) data points. Black lines represent the absolute differences between the real and synthetic waveforms.

1

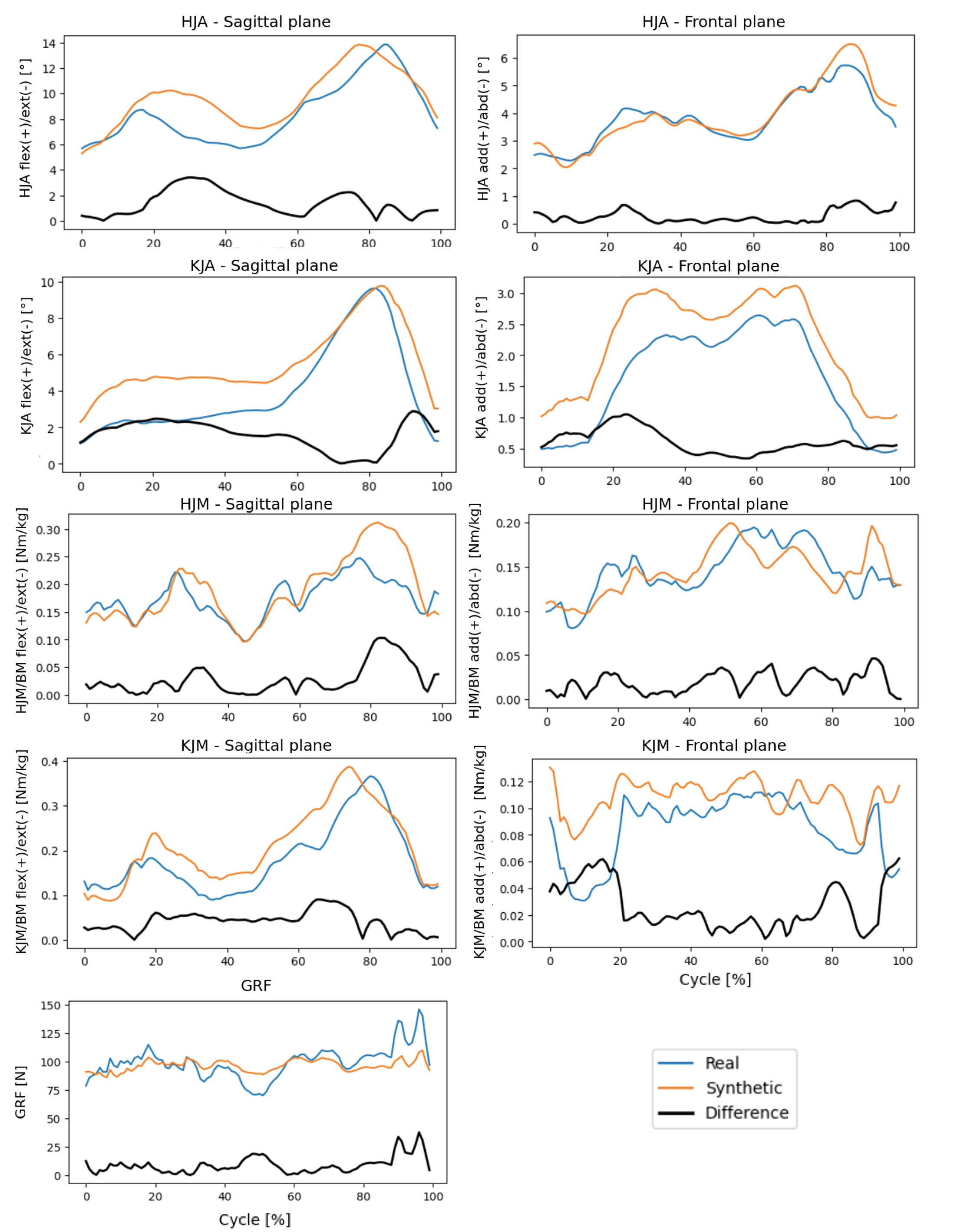

Figure A.7: Graphs showing standard deviation waveforms of all the biomechanical vari- ables analyzed for patients. At each time step, standard deviations were calculated across all corresponding real (blue) and synthetic (orange) data points. Black lines represent the absolute differences between the real and synthetic waveforms.

2
